# Removing weekly administrative noise in the daily count of COVID-19 new cases. Application to the computation of Rt

**DOI:** 10.1101/2020.11.16.20232405

**Authors:** Luis Alvarez, Miguel Colom, Jean-Michel Morel

**Affiliations:** CTIM. Departamento de Informática y Sistemas, Universidad de Las Palmas de Gran Canaria. Spain; Université Paris-Saclay, ENS Paris-Saclay, France

**Keywords:** COVID-19, Weekly administrative noise, Effective Reproduction number, reproduction rate, R0, Rt, SARS-CoV-2, Serial Interval

## Abstract

The way each country counts and reports the incident cases of SARS-CoV-2 infections is strongly affected by the “weekend effect”. During the weekend, fewer tests are carried out and there is a delay in the registration of cases. This introduces an “administrative noise” that can strongly disturb the calculation of trend estimators such as the effective reproduction number *R*(*t*). In this work we propose a procedure to correct the incidence curve and obtain a better fit between the number of infected and the one expected using the renewal equation. The classic way to deal with the administrative noise is to invoke its weekly period and therefore to filter the incidence curve by a seven days sliding mean. Yet this has three drawbacks: the first one is a loss of resolution. The second one is that a 7-day mean filter hinders the estimate of the effective reproduction number *R*(*t*) in the last three days before present. The third drawback of a mean filter is that it implicitly assumes the administrative noise to be additive and time invariant. The present study supports the idea that the administrative is better dealt with as being both periodic and multiplicative. The simple method that derives from these assumptions amount to multiplying the number of infected by a correcting factor which depends on the day of the week. This correcting factor is estimated from the incidence curve itself. The validity of the method is demonstrated by its positive impact on the accuracy of an the estimates of *R*(*t*). To exemplify the advantages of the multiplicative periodic correction, we apply it to Sweden, Germany, France and Spain. We observe that the estimated administrative noise is country dependent, and that the proposed strategy manages to reduce it noise considerably. An implementation of this technique is available at www.ipol.im/ern, where it can be tested on the daily incidence curves of an extensive list of states and geographic areas provided by the European Centre for Disease Prevention and Control.

---

The effective reproduction number *R*(*t*) is one of the most important epidemiological characteristics of the COVID-19 pandemic. It is constantly invoked by politicians and their scientific advisers to steer the social distancing measures. *R*(*t*) represents the expected number of secondary cases produced by a primary case at each time *t*. It can be computed from the incidence curve *i*(*t*) and the serial interval Φ, which is the empirical probability distribution of the time between the onset of symptoms in a primary case and the onset of symptoms in secondary cases. There are different strategies to estimate *R*(*t*) from *i*(*t*) and Φ: EpiEstim, the method proposed in [4] is widely used. In the online interface available at www.ipol.im/ern, we compare our estimate of *R*(*t*) with the one of EpiEstim. In [11] a technique to compute *R*(*t*) separating local transmission and imported cases is proposed. In [6] the authors make a detailed comparison of the methods proposed in [4], [2] and [13]. A systematic review and analysis of the serial interval is presented in [12]. In [10], [5], [7] different statistical distributions of the serial interval Φ for the SARS-CoV-2 are proposed.

The so-called renewal equation (see [9]) is a key epidemiological model linking the daily count of new detected cases of infections, *i*(*t*), with the reproductive power, *A*(*t, s*), at time *t* and infection-age *s* at which an infected individual generates secondary cases. *A*(*t, s*) depends on *R*(*t*) and Φ(*s*) and the renewal equation can be expressed as

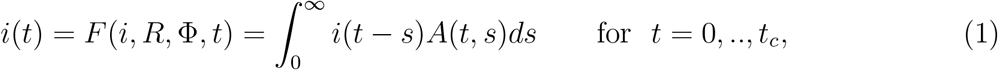

where *t*_*c*_ represents the current time (the last time at which *i*(*t*) is available). For instance, in [4] and [3] the following formulation of the renewal equation is used to compute *R*(*t*) from *i*(*t*) and Φ(*s*):

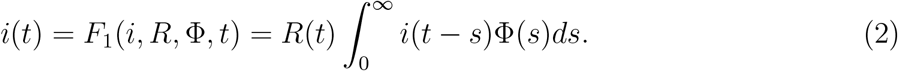

This model is a simplification of the Nishiura equation [8]:

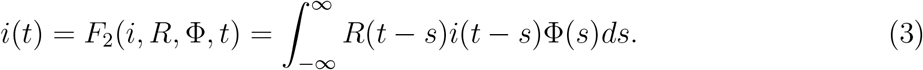

In the original formulation of this model it was assumed that Φ(*s*) = 0 for *s* ≤ 0. However, in the case of SARS-CoV-2 a patient can show symptoms before the patient who infected him/her shows symptoms him/herself. This means that, actually, Φ(*s*) can be positive for *s <* 0. In [1], the above model is used without any restriction on the support of Φ(*s*). Since measurements are generally made daily, a discrete formulation of model (3) is sound. It can be expressed as

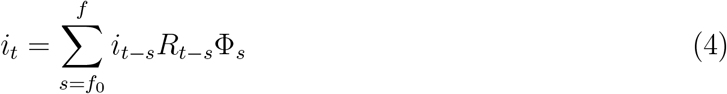

where *f*_0_ is, in general, negative.

In the technique we propose in this work, either of the above expressions of the renewal equation can be used. In the experiments presented, we use the model *F*_2_(*i, R*, Φ, *t*), and the method developed in [1] to estimate *R*_*t*_ from *i*_*t*_ and Φ_*s*_. This method is based on the the minimization of the following energy:

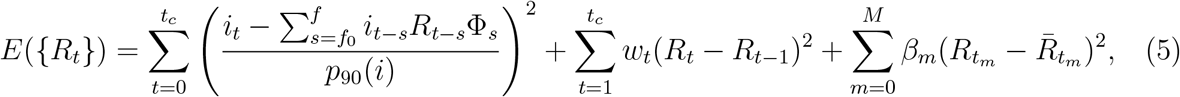

where *p*_90_(*i*) is the 90th percentile of 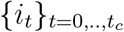 used to normalize the energy with respect to the size of *i*_*t*_. The first term of *E* is a data adjustment term which forces the renewal equation (4) to be satisfied as much as possible. The second term forces *R*_*t*_ to be a smooth curve; *w*_*t*_ *≥* 0 represents the weight of the regularization at each time *t*. The higher the value of *w*_*t*_ the smoother *R*_*t*_. The last term of *E* forces 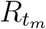 to be close to an initial estimate given by 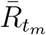 for some particular times *t*_*m*_. Roughly speaking, minimizing the energy *E* leads to satisfy approximately the renewal equation (4) with a reasonably smooth *R*_*t*_ and, optionally, prescribed initial values for some particular times *t*_*m*_.

The daily number of new detected cases, *i*_*t*_, is strongly affected by the “weekend effect”. During the weekend, fewer tests are carried out and there is a delay in the registration of cases. It follows that the actual number of cases is systematically underestimated in some days of the week and overestimated in others. The usual way to deal with this weekly administrative noise is to use a 7-day moving average of *i*_*t*_, but this procedure negatively affects the accuracy of the point estimate of the trend of *i*_*t*_ when approaching the current date and forces stopping the estimate three days before present.

The main assumption of this paper is that a significant part of the discrepancy between *i*_*t*_ and its expected value *F* (*i, R*, Φ, *t*) is given by the weekend effect. We therefore assume that the quotient

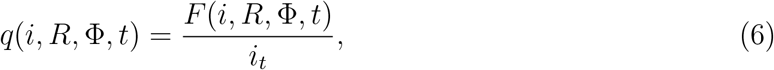

follows a 7-day periodic-dynamic, that is, *q*(*i, R*, Φ, *t*) ≈ *q*(*i, R*, Φ, *t* − 7). In other words, the ratio between *F* (*i, R*, Φ, *t*) and *i*_*t*_ depends mainly on the day of the week. In the experiments shown in this article, it will be observed that, indeed, *q*(*i, R*, Φ, *t*) follows accurately this periodic pattern in several countries^1^. For example, in Sweden every Monday the registered value *i*_*t*_ is systematically underestimated and on Saturdays the opposite effect occurs. So we propose to approximate *q*(*i, R*, Φ, *t*) using a 7-day periodic function given by the vector 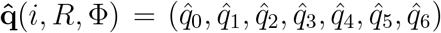. Therefore 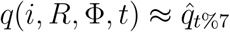 where the symbol % represents the modulo operation, that is, the remainder of the division between two numbers.

To compute 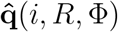 we proceed to a least square estimate

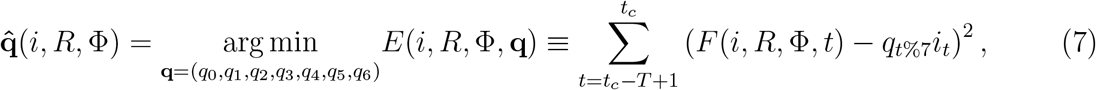

where *T* represents the number of days used in the estimation (in our experiments we use *T* = 56, that is 8 weeks). We point out that the value 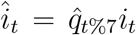 can be considered as an update of *i*_*t*_ where we have removed the weekly administrative noise. To preserve the number of accumulated cases in the period of estimation, we add to the minimization problem (7) the constraint

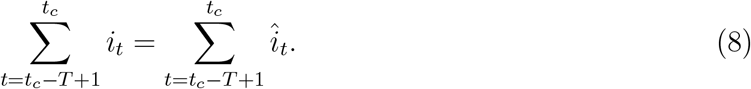

In that way, the multiplication by the factor 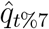 produces a redistribution of the cases *i*_*t*_ during the period of estimation, but it does not change the global amount of cases. Notice that we could add weights to some particular days in the expression of *E*(*i, R*, Φ, **q**) in (7). For example, if a day is a holiday in the middle of the week, one might reduce, in the energy *E*(*i, R*, Φ, **q**), the weight of that day and the following ones.

Once *i*_*t*_ is updated using 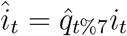, we can use *î*_*t*_ to recompute *R*_*t*_ using the renewal equation (1). We denote by 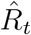 the updated version of *R*_*t*_. This whole procedure is repeated to update iteratively *î*_*t*_ and 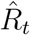 until convergence. The final value *î*_*t*_ and 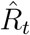 provides a more realistic trend of the evolution of *i*_*t*_ and improves the estimation of *R*_*t*_. The final vector 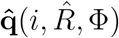 represents the set of multiplicative factors used to remove the weekly administrative noise. In Fig. 1 we present a flowchart of the whole procedure and in the appendix we give technical details.

**Figure 1:**
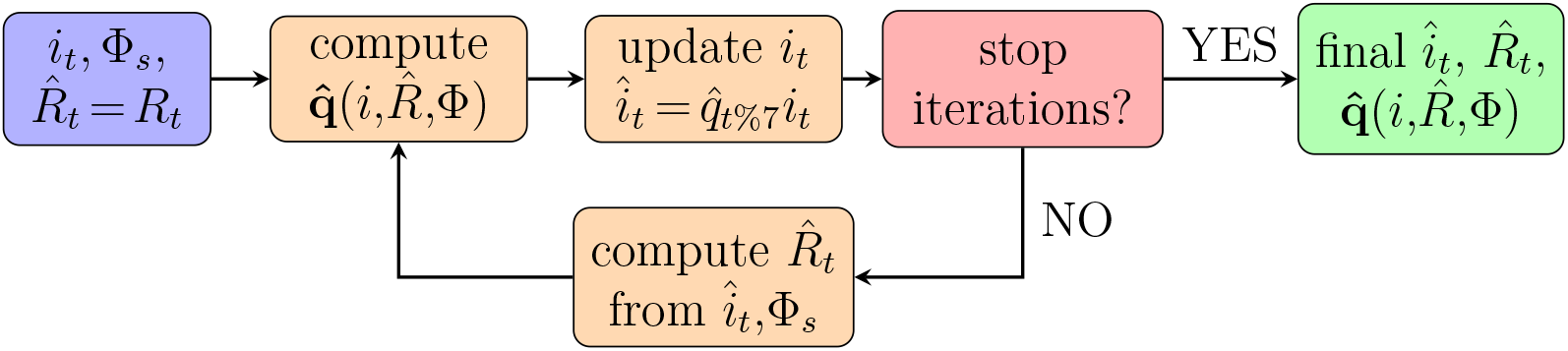
Flowchart of the estimation of 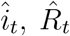 and 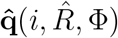. First *R*_*t*_ is computed from *i*_*t*_ and Φ_*s*_ using the method proposed in [1] and 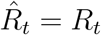 is initialized. Next, 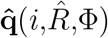 is obtained by minimizing (7) with the constraint (8). Then *i*_*t*_ is updated as 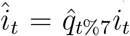. The iteration is stopped when the efficiency measure ℐ defined in formula (9) does not improve in the current iteration. Otherwise, 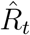 is updated by the method proposed in [1] from *î*_*t*_ and Φ_*s*_ and the iteration goes on.

## RESULTS

All of the experiments made here can be reproduced with the online interface available at www.ipol.im/ern. In the appendix, we have included some details about the use of this online demo. To measure how well the removal of the weekly administrative noise improves the explanation of *i*_*t*_ by the renewal equation *F* (*i, R*, Φ, *t*), we use the following efficiency measure:

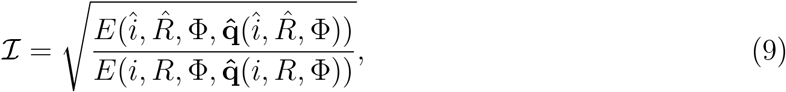

where *E*(.) is defined in (7). ℐ represents the reduction, after the removal of the weekly administrative noise, of the average distance between *i*_*t*_ and *F* (*i, R*, Φ, *t*). The smaller ℐ, the more efficient the noise reduction has been. In fact, the value of ℐ can be used to assess whether it is worth applying the proposed method to a given country and in a given time interval.

In Fig. 2 we plot, for Sweden, Germany, France and Spain the values of the vector 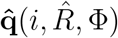 obtained for each day of the week. We observe that in Sweden and Germany, Monday is the day of the week where the value of *i*_*t*_ is most underestimated (the higher 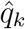 is, the more underestimated is *i*_*t*_ in the day *k*). However, in France, that day corresponds to Tuesday. This suggests that in France there is an additional delay of one day due to the way France records and reports the number of new cases. In these three countries, the effect of weekly administrative noise is mainly concentrated on Monday, Tuesday and Wednesday (with a 1-day lag in the case of France). The case of Spain is special because it does not provide data on the weekend. In that case we have assumed that on Saturday, Sunday and Monday the number of cases is constant and equal to the accumulated number of cases on Saturday, Sunday and Monday divided by 3. Moreover, in Spain, some regions sometimes do not report data for one or several days, which produces an additional administrative noise different from the weekend effect. The more orderly the daily count of new infected, the better the result obtained by our method.

**Figure 2:**
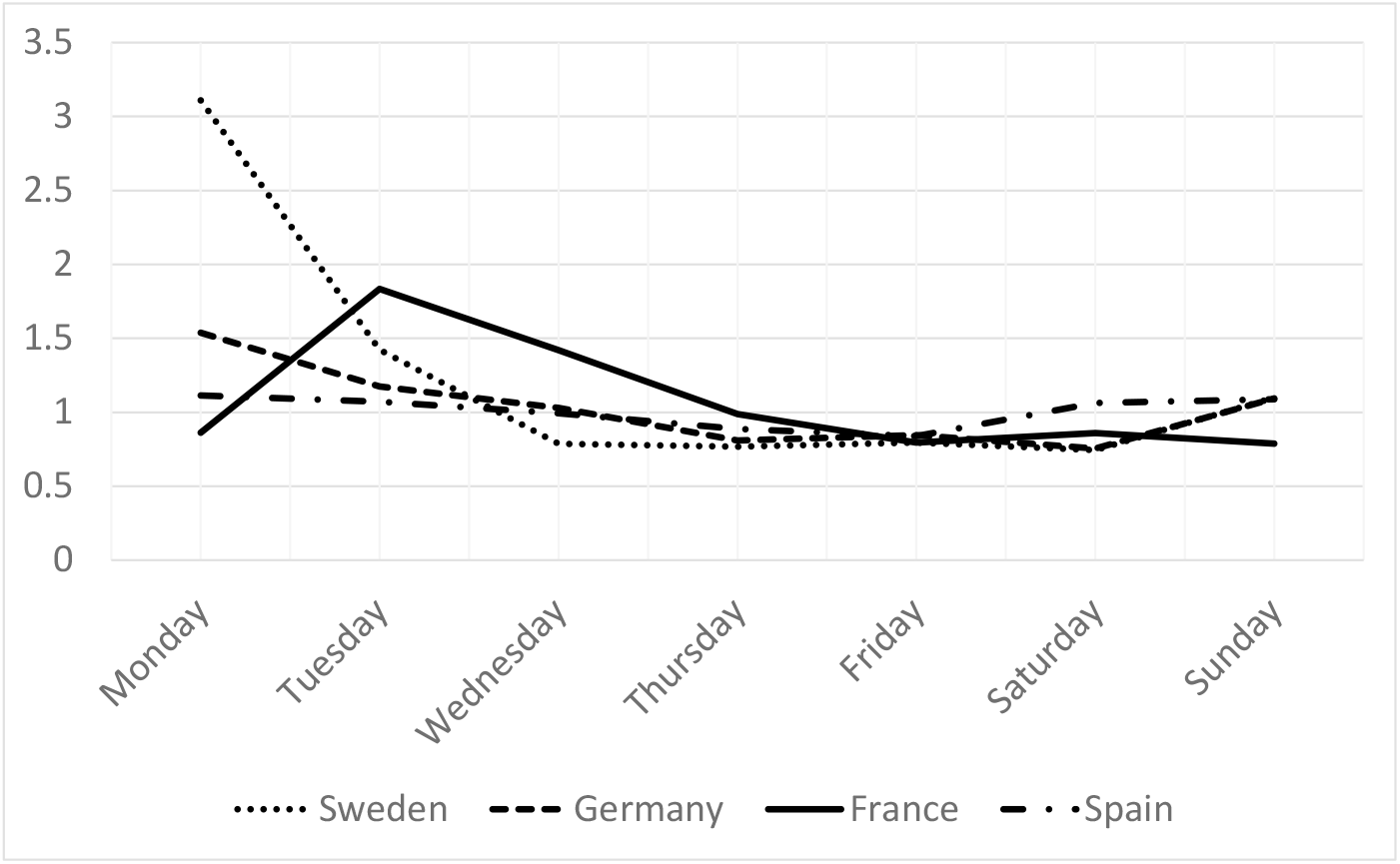
Plot of 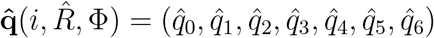 for different countries.

In table 1 we present the numerical values of the vector 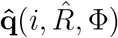, the efficiency measure ℐ and the effective reproduction number in the current time before and after the weekly administrative noise removal given by 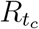 and 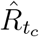 respectively. Notably, and with the exception of Spain, the efficiency value ℐ shows a high reduction of the administrative noise. At the end of the period chosen for the study, between September 9 and October 28, there is a strong expansion of the virus. This period ends on a Wednesday. In the previous days the number of cases was underestimated. It follows that the estimated effective reproduction number becomes considerably higher after eliminating the weekly administrative noise.

**Table 1:** We present for Sweden, Germany, France and Spain the values of the vector 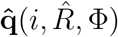 for each day of the week, the efficiency measure, ℐ, and the effective reproduction number in the current time before and after the administrative noise removal given by 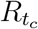 and 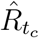 respectively.

|  | Sweden | Germany | France | Spain |
| --- | --- | --- | --- | --- |
| Monday ($\hat{q}_0$) | 3.109167 | 1.537838 | 0.863379 | 1.113635 |
| Tuesday ($\hat{q}_1$) | 1.424005 | 1.175497 | 1.834212 | 1.071432 |
| Wednesday ($\hat{q}_2$) | 0.787374 | 1.029281 | 1.419457 | 0.993049 |
| Thursday ($\hat{q}_3$) | 0.766827 | 0.808254 | 0.989091 | 0.887398 |
| Friday ($\hat{q}_4$) | 0.795438 | 0.846798 | 0.794311 | 0.838609 |
| Saturday ($\hat{q}_5$) | 0.744447 | 0.754752 | 0.858157 | 1.063243 |
| Sunday ($\hat{q}_6$) | 1.100563 | 1.093202 | 0.788024 | 1.087866 |
| $\mathcal{I}$ | 0.2526414 | 0.2717397 | 0.3725009 | 0.6439962 |
| $R_{t_c}$ | 1.426663 | 1.470402 | 1.277142 | 1.225078 |
| $\hat{R}_{t_c}$ | 1.509397 | 1.637230 | 1.563844 | 1.263821 |

In Fig. 3 - 6 we plot on the one hand the quotient 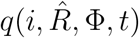 and its periodic approximation 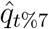, and on the other hand the values of *i*_*t*_, its update *î*_*t*_ after the removal of the weekly administrative noise and the expected value using 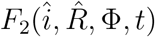. For Sweden, Germany and France we observe a quite a good agreement between the quotient 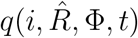 and its periodic approximation 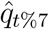, which supports the validity of the proposed method and our assumption that the evolution of 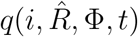 follows a 7-day periodic dynamic. For these countries, 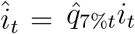 the number of new cases after the removal of the administrative noise, is less oscillating than *i*_*t*_ and very close to its expected value following the renewal equation 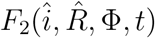. In the case of Spain, the obtained improvement is minor. On the one hand, Spain does not report data during the weekend, which introduces an additional disturbance in the data that hinders the application of the proposed method. In addition, in two of the weeks included in the period of the study, 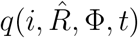 diverges significantly from its periodic approximation 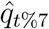.

**Figure 3:**
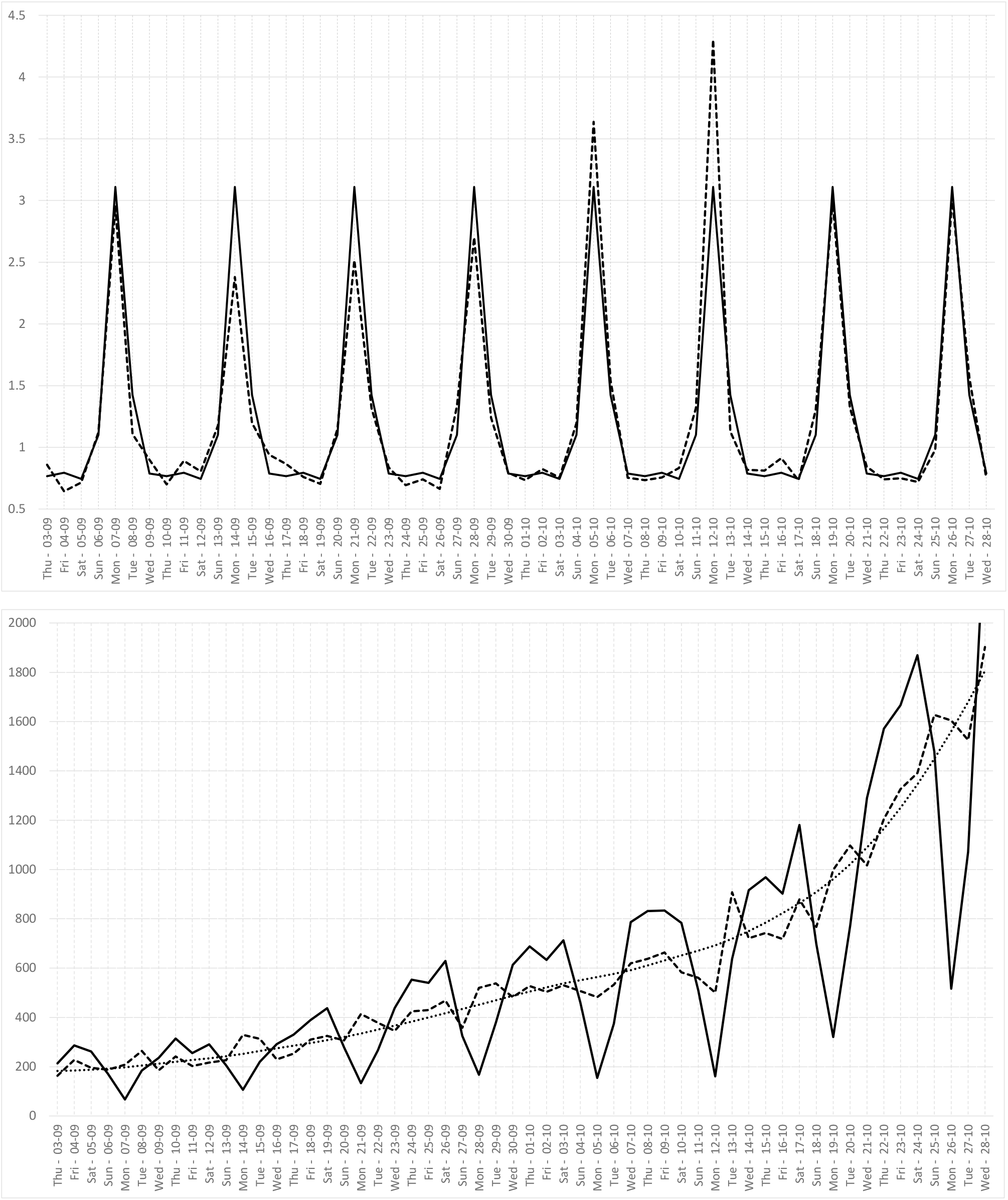
In the case of Sweden, we plot, between September 9 and October 28: **(up)** 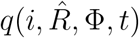 (dotted line) and its periodic approximation 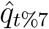 (solid line). (**down**): *i*_*t*_ (solid line), its update *î*_*t*_ (dashed line) removing the administrative noise and the expected value using 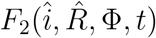 (dotted line).

**Figure 4:**
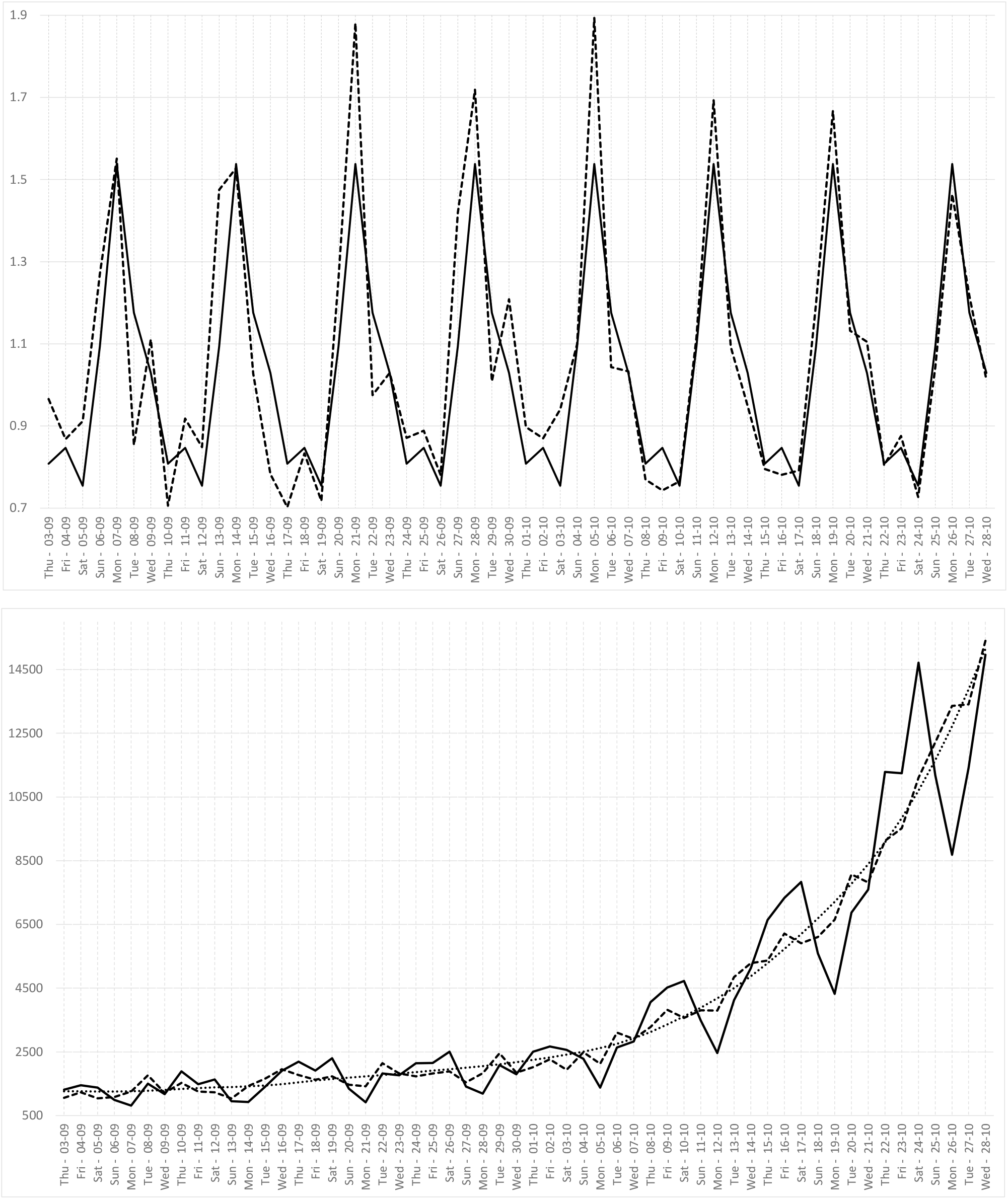
In the case of Germany, we plot, between September 9 and October 28: **(up)** 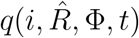 (dotted line) and its periodic approximation 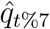 (solid line). (**down**): *i*_*t*_ (solid line), its update *î*_*t*_ (dashed line) removing the administrative noise and the expected value using 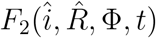 (dotted line).

**Figure 5:**
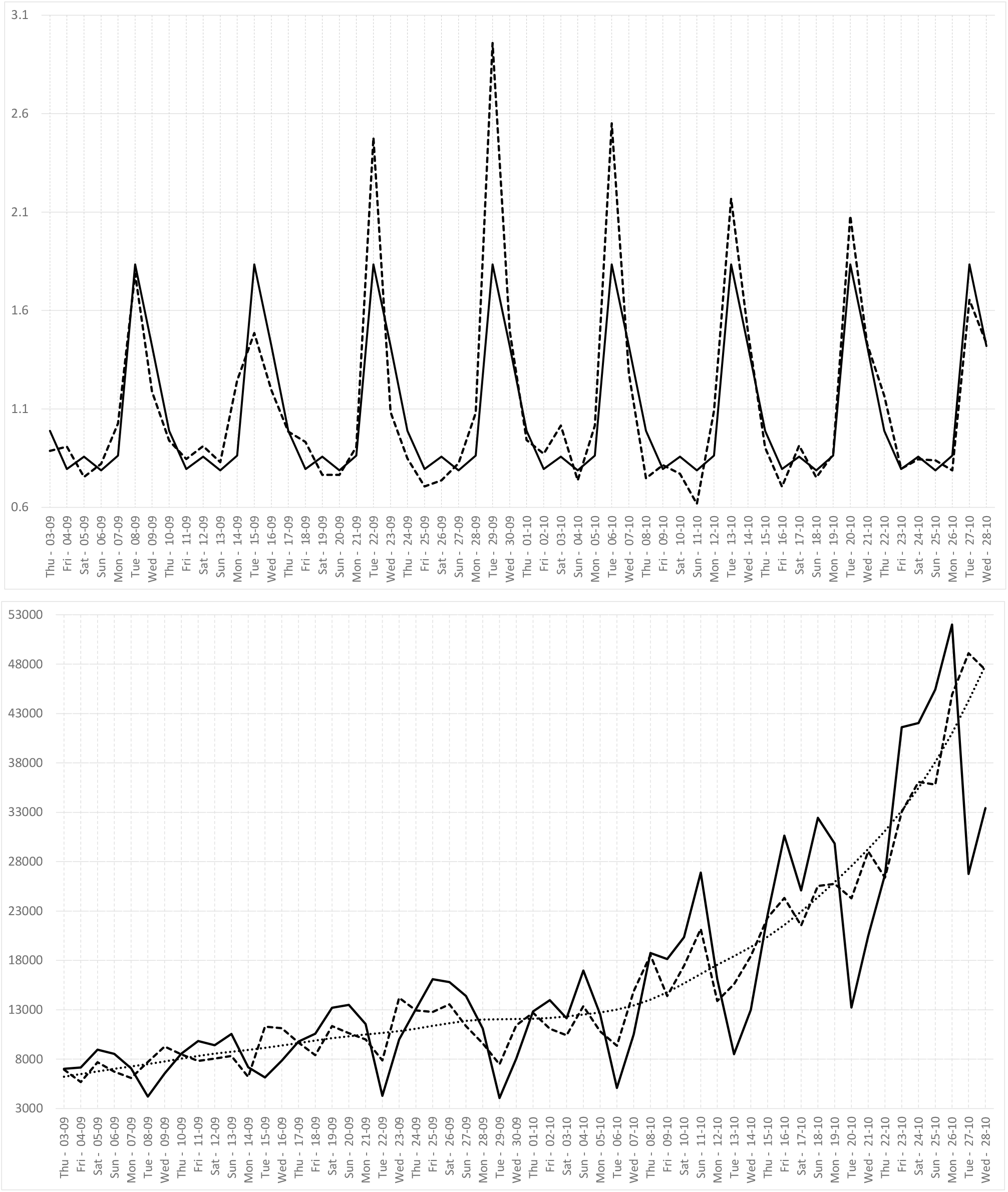
In the case of France, we plot, between September 9 and October 28: **(up)** 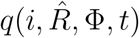 (dotted line) and its periodic approximation 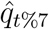 (solid line). (**down**): *i*_*t*_ (solid line), its update *î*_*t*_ (dashed line) removing the administrative noise and the expected value using 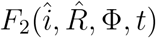 (dotted line).

**Figure 6:**
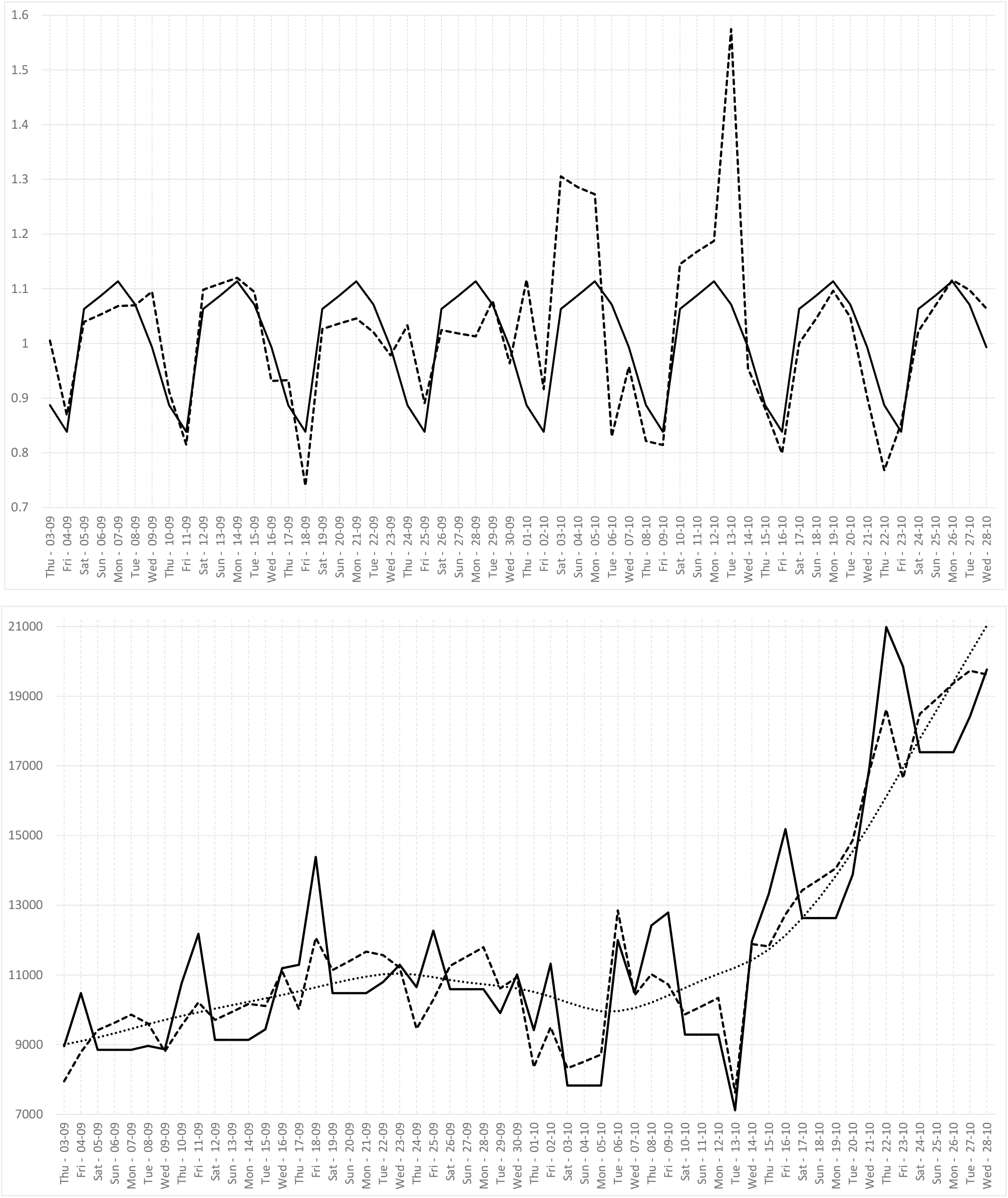
In the case of Spain, we plot, between September 9 and October 28: **(up)** 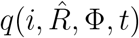 (dotted line) and its periodic approximation 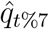 (solid line). (**down**): *i*_*t*_ (solid line) its update *î*_*t*_ (dashed line) removing the administrative noise and the expected value using 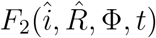 (dotted line).

## 1 CONCLUSION

We proposed a technique to remove the weekly administrative noise in the incidence curves of COVID-19, based on the improvement of the agreement between *i*_*t*_ and its expected value using the renewal equation *F* (*i, R*, Φ, *t*). The method boils down to multiplying *i*_*t*_ by a factor 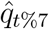 which depends on the day of the week. The main assumption supporting the validity of this approach is that the quotient of *i*_*t*_ and *F* (*i, R*, Φ, *t*) follows approximately a 7-day periodic dynamic. We verified this assumption on Sweden, Germany and France. In the case of Spain the method brings less improvement, due to the fact that Spain does not report data on weekends. The proposed method updates iteratively *i*_*t*_ and the effective reproduction number *R*_*t*_. The number of cases after the removal of the weekly administrative noise 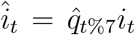 is much less oscillating than *i*_*t*_ and very close to its expected value following the used renewal equation. From *î*_*t*_ we obtain a more accurate value of the effective reproduction number *R*_*t*_. An implementation of this technique is available at www.ipol.im/ern. In this online interface we compare the final estimation of *R*_*t*_ with the estimate obtained by EpiEstim. As shown in [3], in practice, EpiEstim uses a 7-day moving average to remove the administrative weekly noise. As can be observed using the online interface, this 7-day moving average introduces a shift towards the past that is not observed in our estimate. Therefore our method seems to provide a more to date estimate of *R*_*t*_ than EpiEstim.

## Data Availability

All data referred to in the manuscript are publicly available at the European Centre for Disease Prevention and Control.

https://www.ipol.im/ern

## Abbreviations

EpiEstim: Software to compute the effective reproduction number proposed by Cori et al. in the paper: *A new framework and software to estimate time-varying reproduction numbers during epidemics* published in the American Journal of Epidemiology.
*R*(*t*): Effective Reproduction Number. To differentiate between the continuous and discrete cases, we use the notation *R*(*t*) in the continuous case and *R*_*t*_ in the discrete case.
*i*(*t*): incidence curve, the number of daily tested positive registered. To differentiate between the continuous and discrete cases, we use the notation *i*(*t*) in the continuous case and *i*_*t*_ in the discrete case.
Φ: serial interval.

## Appendix

### Technical issues

Here, we show some technical details of the proposed technique. First we notice that, in general, the only data we use is *i*_*t*_ and Φ_*s*_. So initially, we do not know the actual day of the week which corresponds to each datum *i*_*t*_. For the computation of 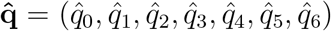, we will assume, without loss of generality, that the first day of the period we consider for the estimation, that is *t*_*c*_ *− T* + 1 corresponds to the time *t* = 0. This means that we are initially assuming that this day corresponds to a Monday, but in fact, this is not relevant because we can reorganize at any time the values of 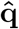 to fit the actual days of the week.

The minimization problem (7) given by the quadratic energy:

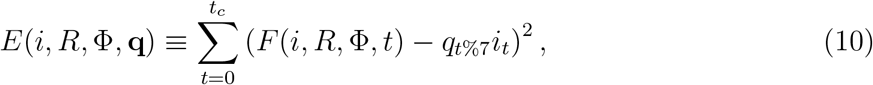

can be expressed in matrix form as

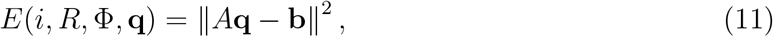

where ‖.‖ is the usual Euclidean norm, *A ∈ R*^*T×*7^ is defined by

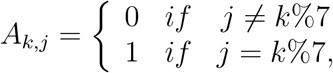

**b** ∈ *R*^*T*^ is defined by *b*_*k*_ = *F* (*i, R*, Φ, *k*). It is well-known that the above minimization problem has a closed form solution given by

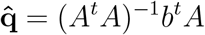

The constraint (8) given by

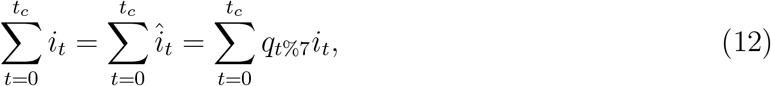

can be expressed as the following additional linear equation:

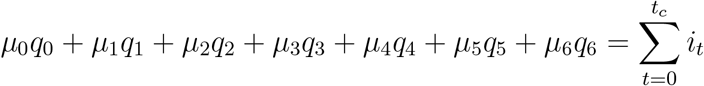

where

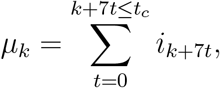

this constraint can be included in the minimization procedure by removing one of the unknowns using the above equation. However, we use another approach which consists of adding the above equation, multiplied by a certain weight *w*, to the expression (11). In this way we can control the weight that is assigned to this constraint. If *w* is large, we force the restriction to be fulfilled and if *w* = 0 we remove this constraint. In the experiments performed in this work we use *w* = 10^5^, so we choose that the constraint is satisfied.

As explained in the Fig. 1, in the proposed method, we update iteratively *i*_*t*_, *R*_*t*_ and 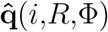. Let us denote by 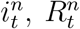 and 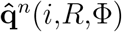 these updates for each iteration *n* starting at *n* = 0. Following the flowchart of Fig. 1 these updates are computed using the following algorithm:

#### Algorithm 1: Algorithm to estimate 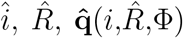. *MaxIter* represents the maximum number of iterations allowed.

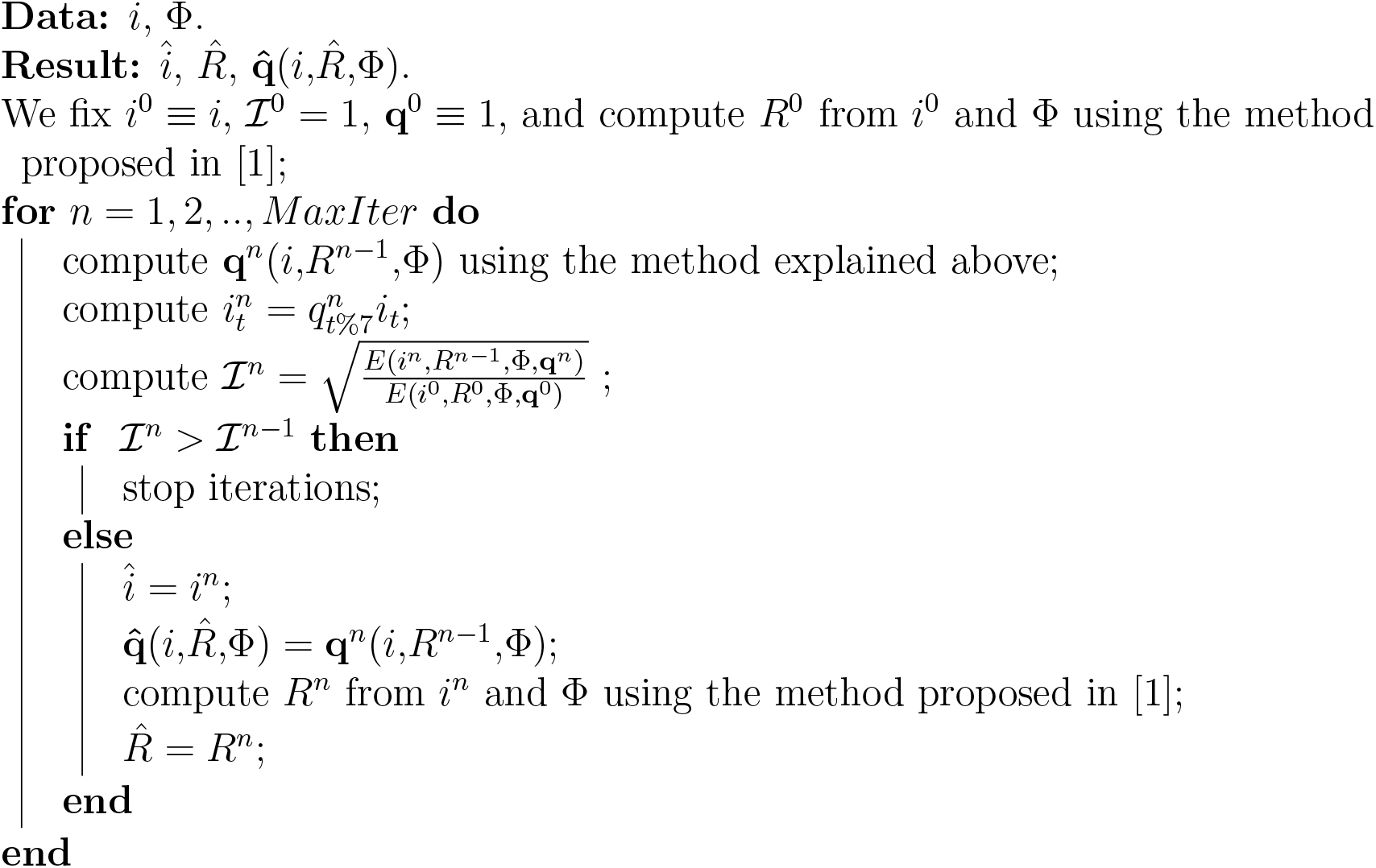

### Using the online interface

In the online interface, available at www.ipol.im/ern, one can test the method proposed in [1] to estimate *R*_*t*_ as well as the method proposed here to remove the weekly administrative noise. When the DEMO is executed with the option “remove weekly administrative noise” activated, the method proposed in this paper is used to compute *î*_*t*_, 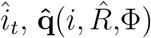 and 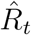. A comparison with the EpiEstim method is showed. We also show (in the plot on the right) the original curve *i*_*t*_ as well as the estimation using the renewal equation 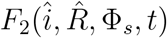. In the output file named “Rn.csv” you will find in the first row, the final value of the efficiency measure ℐ (see (9)) and the vector 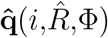. The elements of the vector are organized in such a way that the last one 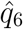 corresponds to the multiplicative factor of the last element of *i*_*t*_ (that is 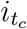). In this way, to obtain *î*_*t*_ we have just to do 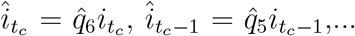 and, in general: 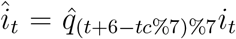. Moreover, in the first column of the output file “Rn.csv” you will find the values of 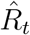, in the second column, 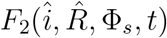, in the third column, the original number of infected *i*_*t*_, and in the forth column a measure of the variability in the estimation of 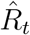, as explained in [1].

## Footnotes

1 To test this assumption on many more countries, the reader is invited to the online demo www.ipol.im/ern

